# Multidimensional analysis of immune response identified biomarkers of recent *Mycobacterium tuberculosis* infection

**DOI:** 10.1101/2021.01.27.21250605

**Authors:** Tessa Lloyd, Pia Steigler, Cheleka A.M. Mpande, Virginie Rozot, Boitumelo Mosito, Constance Shreuder, Timothy D. Reid, Mark Hatherill, Thomas J. Scriba, Francesca Little, Elisa Nemes, the ACS Study Team

## Abstract

The risk of tuberculosis (TB) disease is higher in individuals with recent *Mycobacterium tuberculosis* (*M*.*tb*) infection compared to individuals with more remote, established infection. We aimed to define blood-based biomarkers to distinguish between recent and remote infection, which would allow targeting of recently infected individuals for preventive TB treatment. We hypothesized that integration of multiple immune measurements would outperform the diagnostic performance of a single biomarker. Analysis was performed on different components of the immune system, including adaptive and innate responses to mycobacteria, measured on recently and remotely *M*.*tb* infected adolescents. The datasets were standardized using variance stabilizing (vast) scaling and missing values were imputed using a multiple factor analysis-based approach. For data integration, we compared the performance of a Multiple Tuning Parameter Elastic Net (MTP-EN) to a standard EN model, which was built to the single datasets. Biomarkers with non-zero coefficients from the optimal single data EN models were then isolated to build logistic regression models. A decision tree and random forest model were used for statistical validation. We found no difference in the predictive performances of the optimal MTP-EN model and the EN model [average area under the receiver operating curve (AUROC)=0.93]. EN models built to the integrated dataset and the adaptive dataset yielded identically high AUROC values (average AUROC=0.91), while the innate data EN model performed poorly (average AUROC=0.62). Results also indicated that integration of adaptive and innate biomarkers did not outperform the adaptive biomarkers alone (Likelihood Ratio Test *χ*^2^=6.09, p=0.808). From a total of 193 variables, the level of HLA-DR on ESAT6/CFP10-specific Th1 cytokine-expressing CD4 cells was the strongest biomarker for recent *M*.*tb* infection. The discriminatory ability of this variable was confirmed in both tree-based models.

A single biomarker measuring *M*.*tb*-specific T cell activation yielded excellent diagnostic potential to distinguish between recent and remote *M*.*tb* infection.

## 1 Introduction

Tuberculosis (TB) is an airborne bacterial disease that is a leading cause of mortality due to an infectious agent worldwide [1]. It is estimated that about a quarter of the world’s population is infected with *Mycobacterium tuberculosis* (*M*.*tb*), the causative agent of TB [2]. Acquisition of *M*.*tb* infection is generally asymptomatic and often remains undiagnosed unless serial diagnostic testing is performed. To determine *M*.*tb* infection status, the QuantiFERON TB (QFT) measures the level of interferon-gamma (IFN-*γ*), a cytokine released by T cells, upon stimulation of blood cells with two immunodominant antigens expressed by *M*.*tb*, early secretory antigen 6 (ESAT6) and culture filtrate protein 10 (CFP10) (here collectively termed E6C10). The highest risk of progressing to TB disease is during the first two years post-infection [3], which can be measured as recent QFT conversion by serial testing. However, serial testing for *M*.*tb* infection is not routinely performed in TB endemic settings. A blood-based immune signature that enables identification of recent *M*.*tb* infection would therefore allow targeting of preventive treatment to those at high risk of TB progression, even without serial diagnostic testing.

In order to define immunological determinants of recent *M*.*tb* infection, data from different arms of the immune response, namely adaptive, donor unrestricted T (DURT) and innate cell immunity were combined. Adaptive immunity consists of memory-driven antigen-specific T cell responses, such as those measured by QFT. In this study we measured functional and phenotypic features of classical *M*.*tb*-specific T cell responses and refer to these variables as the adaptive dataset. In contrast, innate immune cells, such as monocytes or natural killer (NK) cells, provide non-specific cellular defence mechanisms, which are more transient in nature. DURT cells display features of both adaptive and innate immune cells and bridge both arms. In this study, we included measurements of monocyte, NK and DURT cell functions in the innate dataset. We hypothesized that the integration of multiple immune measures from the adaptive and innate immune arms would outperform individual data types in stratifying individuals with recent or remote *M*.*tb* infection.

Data integration presents several challenges, such as: i) different scales from different data types, which is typically overcome by employing data standardization or transformations methods; ii) missing values that arise due to some individuals or time points not being available in each data table, which can be meaningfully replaced using imputation methods; and iii) high dimensionality of the dataset post-integration.

Regularized regression with sparsity is a common approach for modeling high-dimensional datasets with multicollinearity. The most popular regularized regression models are Ridge Regression [4], which minimizes the residual sum of squares subject to an L2 bound; the least absolute shrinkage and selection operator (LASSO) model [5], which imposes an L1 penalty on the regression coefficients; and the Elastic Net (EN) model [6], which is a combination of the two. The latter two models are particularly advantageous as they perform both parameter estimation and feature selection simultaneously, by shrinking the effect of some coefficients to zero. However, if there is a group of highly correlated variables in the dataset, the LASSO model will select one of these variables at random and ignore the rest. The EN model was designed to overcome this issue. Liu et al. (2018) [7], however, observed that the standard EN approach tends to shrink all features simultaneously and does not consider differing effect sizes in predictors from different datasets. The authors hypothesized that the Multiple Tuning Parameter Elastic Net (MTP-EN) model that allows for different degrees of shrinking for variables from different data sets could account for the differences between each dataset and result in a model with higher predictive performance than a standard EN approach. We therefore tested whether the MTP-EN did improve the predictive performance of the integrated dataset, by directly comparing the MTP-EN and standard EN models.

Tree-based algorithms are a common collection of machine learning classification models, consisting of simple decision trees [8] or the popular random forest (RF) model [9]. A classification decision tree is a supervised model that aims to predict a target by learning decision rules from features in a dataset. Decision trees allow easy interpretation of data clearly ranking the importance of feature and relations between predictors. A downfall, however, is that they suffer from high sampling variability [10]. RF models extend decision trees by building multiple trees on bootstrapped samples of the data and merging them together for making decisions to achieve stable and accurate predictions. RF models also introduce additional randomness by considering a random subset of m¡p predictor variables, where p is the total number of predictors in the dataset, as potential split candidates. The importance of each predictor variable can then be quantified by averaging the total amount by which the Gini Index, a measure of node homogeneity, is decreased for a split over a given predictor over all trees. A large value will be indicative of an important predictor. The misclassification error is a natural measure of performance for the RF model.

Biomarkers identified by the regression models were validated via an internal validation procedure using the tree-based algorithms.

## 2 Results

An overview of the data analysis pipeline is provided in Figure 1.

**Figure 1:**
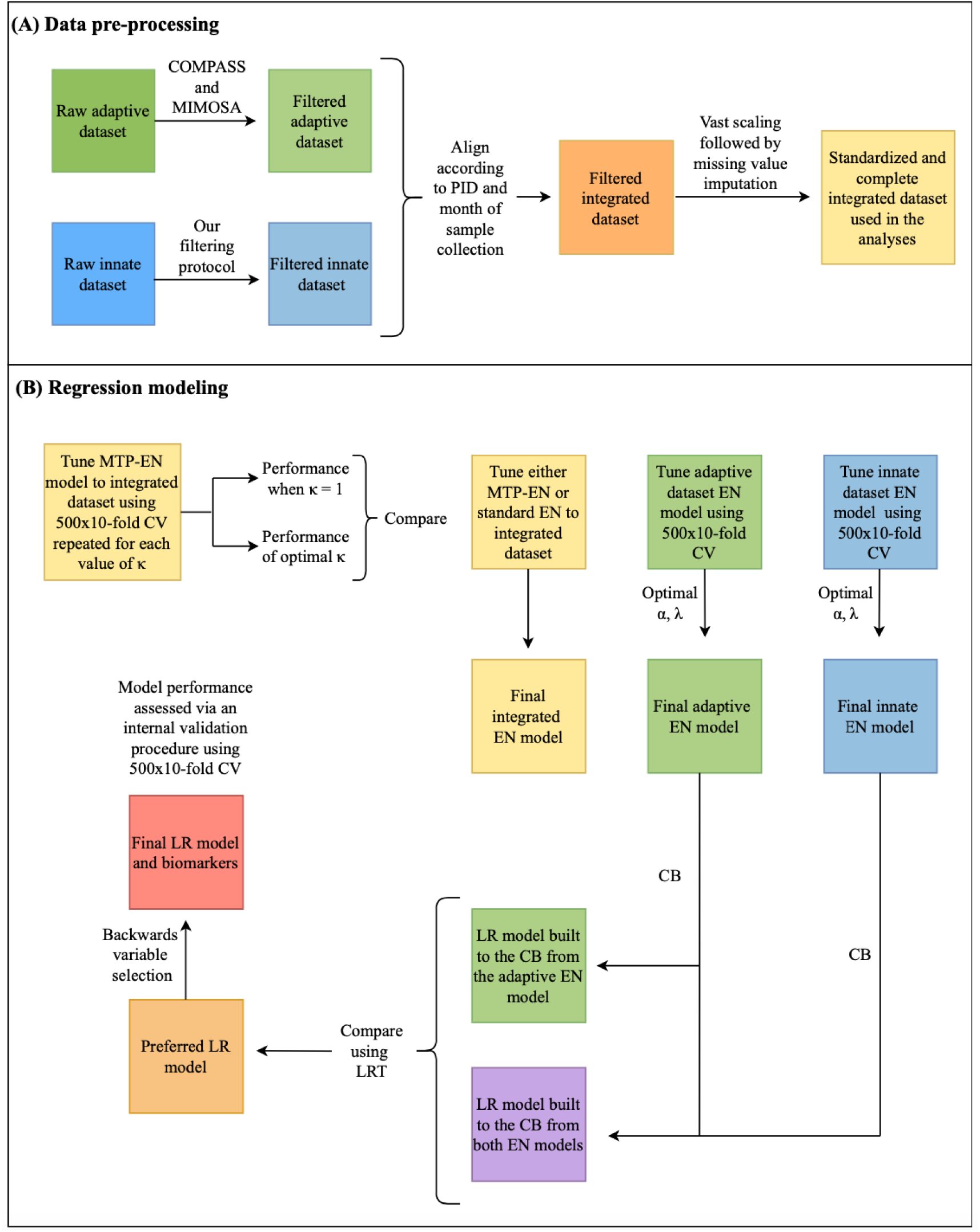
Workflow showing data pre-processing steps (A) and regression modeling (B). PID: participant ID; AUC: area under the curve; LR: logistic regression; CB: candidate biomarkers; CV: cross validation; LRT: likelihood ratio test.

### 2.1 Data pre-processing

In order to successfully integrate the two datasets, several data pre-processing steps needed to be addressed. Data filtering of the adaptive immune response features using COMPASS (Supplementary Figure 1, Supplementary Methods S2.1) and MIMOSA (Supplementary Methods S2.2) retained 132 out of the 259 original variables in the dataset. Further, our novel filtering method (Supplementary Figure 2, Supplementary Methods S2.3) identified 61 biologically meaningful innate features from the 304 variables in the raw dataset. Therefore, among the features considered as biomarkers were 132 functional and phenotypic features of classical *M*.*tb*-specific T cell responses, 6 features of monocytes and 15 features of NK cells. In addition, 12 mucosal associated invariant T (MAIT) cell, 10 gamma-delta (*γδ*) T cell, 6 NKT cell and 12 B cell features were also included, such that a total of 193 variables comprised the filtered, integrated dataset that was used for analyses. We standardized the raw values in this dataset using vast scaling and employed a multiple factor analysis (MFA)-based imputation method [24] to account for missing data points. We found MFA imputation to outperform all other imputation methods tried (Supplementary Figure 3, Supplementary Table 2, Section 5.7).

**Figure 2:**
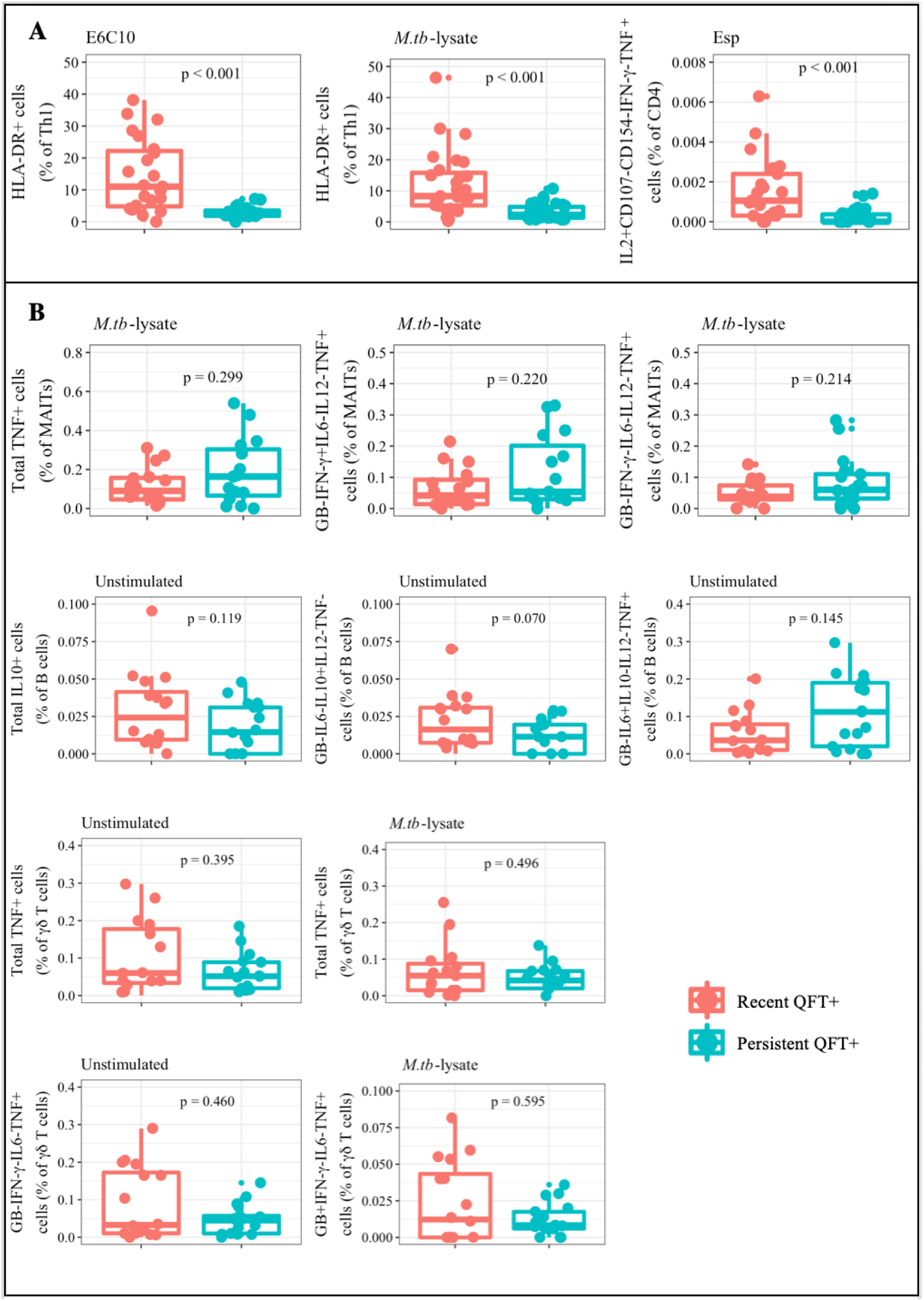
Candidate biomarkers of recent *M.tb* infection identified by the adaptive and innate EN models. Boxplots comparing the raw values of recent (red) and persistent (blue) QFT+ individuals for the three candidate biomarkers identified by the adaptive EN model (A) and the 10 identified by the innate EN model (B). Wilcoxon tests were used to compare the two groups and the resulting p-values are shown.

**Figure 3:**
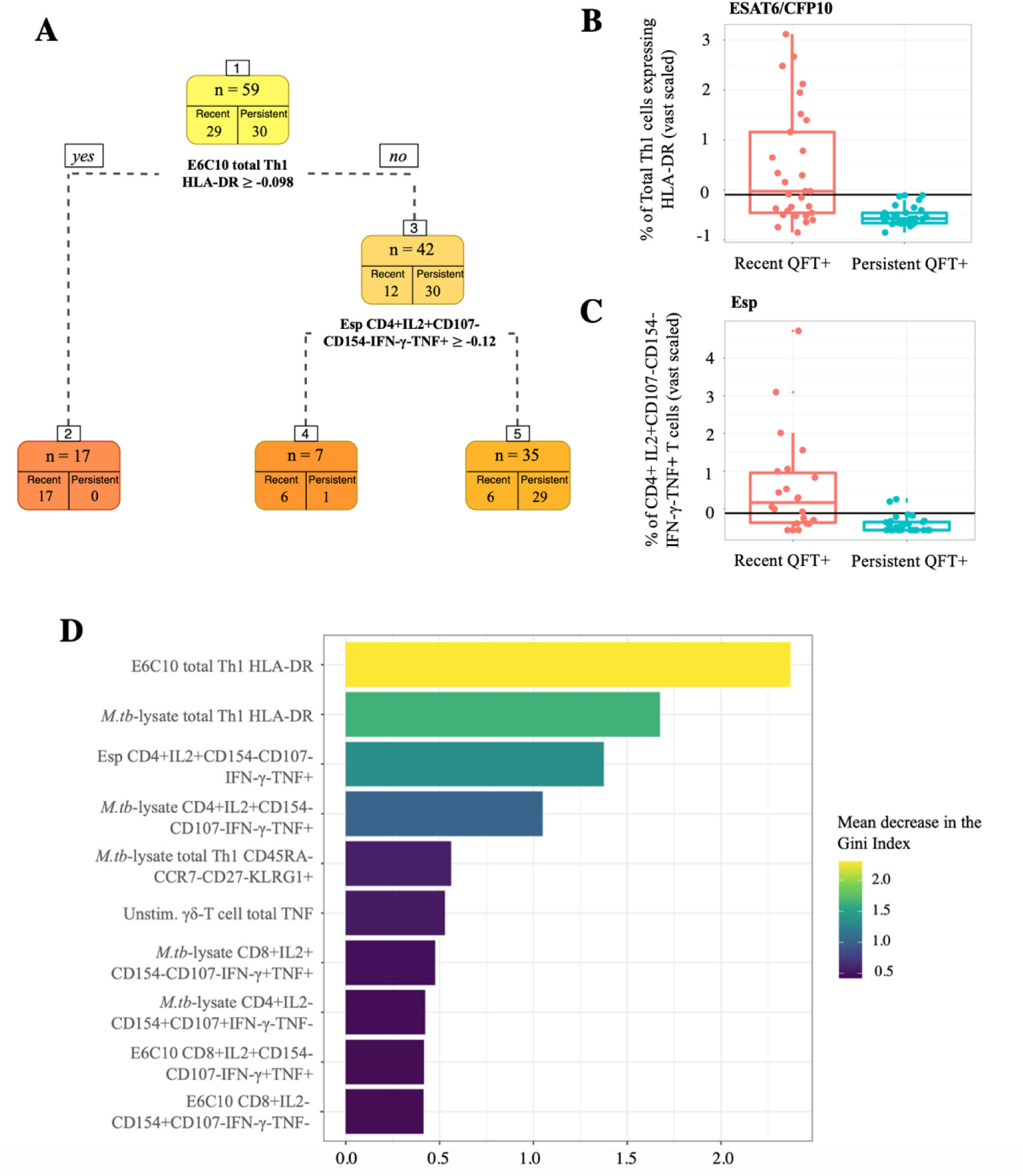
Statistical validation using tree-based models. (A) Results from the simple classification tree built to the entire integrated dataset. Boxplots comparing vast scaled values of recent (red) and persistent (blue) QFT+ individuals were plotted for the two most stratifying features identified by the decision tree. The split values are superimposed onto the plots at (B) −0.098 for proportions of E6C10-specific Th1 cells expressing HLA-DR, and (C) −0.12 for frequencies of Esp-specific IL2+CD107-CD154-IFN-*γ*-TNF+ CD4+ T cells. (D) Variable importance plot of the final RF model showing the top 10 variables that resulted in the largest average decrease in the Gini Index.

### 2.2 The MTP-EN model

The MTP-EN model was built including all 193 variables in the integrated dataset in order to assess whether applying different penalties to each dataset improved the predictive performance of the model, measured by the area under the receiver operating characteristic curve (AUROC) on the testing data. The highest AUROC values after 10-fold cross validation (CV) were stored for each of the 500 repeats, and the performance of the MTP-EN model for each value of *κ*, the ratio of the penalty parameter for the innate dataset relative to that for the adaptive dataset, was reported as an average of the 500 AUROC values. The effect on the testing AUROC for different values of the tuning parameters as measured by *κ* in the MTP-EN is shown in Supplementary 4. The model performance reached a plateau for *κ >* 0.8, where values of *κ <* 1 result in a smaller penalty applied to the innate dataset. Differential penalization therefore resulted in lower comparative AUROC values when the effect sizes of the features in the adaptive dataset in particular were decreased relative to the innate dataset (*λ*_1_ *> λ*_2_). In terms of the predictive performance as determined by the AUROC, the standard EN (*κ* = 1) and the optimal MTP-EN model with *κ* = 1.7 were identical with an average AUROC of 0.93. In addition, the computation time for the MTP-EN model was significantly longer (24.4 hours) compared to the EN model (2.52 hours). Accordingly, for this specific dataset, there was no added benefit to fitting the MTP-EN model over the standard EN model. The EN model was therefore used for subsequent analyses.

### 2.3 Regularized regression, biomarker discovery and validation

EN models were subsequently built to test whether an integrated model outperforms or adds to the single dataset models. Therefore, three EN models were built to the integrated dataset, and the adaptive and the innate data types separately. The final EN models built on the adaptive variables had tuning parameter values of *α*=0.21 and *λ*=0.82 and identified three candidate biomarkers corresponding to an average AUROC value of 0.91 (Supplementary Figure 5A). The biomarkers identified were proportions of total Th1 cells expressing the phenotypic marker human leukocyte antigen (HLA)-DR, identified by stimulation with either E6C10 or *M*.*tb*-lysate, and the frequency of interleukin (IL)2+CD107-CD154-IFN-*γ*-tumor necrosis factor (TNF)+ CD4+ T cells stimulated with EspC, EspF and Rv2348c (collectively termed Esp). Comparing the two groups (recent *versus* persistent QFT+ individuals) using a Wilcoxon non-parametric test [25], values for these variables were found to be significantly higher (p *<* 0.001) in recent compared to persistent QFT+ individuals (Figure 2A). Hence, these variables could be validated as potential biomarkers for recent infection.

For the EN model built on the innate dataset, the optimal values for *α* and *λ* were 0.18 and 0.41 respectively. This model identified 10 candidate biomarkers, which yielded a poor average AUROC of 0.62 (Supplementary Figure 5B). The 10 candidate biomarkers consisted of different functional subsets from of MAIT cells, B cells and *γδ* T cells (Figure 2B). The MAIT cell subsets were all *M*.*tb*-lysate-reactive and included total TNF+ cells, Granzyme-B (GB)-IFN-*γ*-IL6-IL12-TNF+ and GB-IFN-*γ*+IL6-IL12-TNF+ cells. The *M*.*tb*-lysate-reactive *γδ* T cell subsets included total TNF+ cells and GB+IFN-*γ*-IL6-TNF+ cells, while the unstimulated *γδ* T cell subsets were total TNF+ cells and GB-IFN-*γ*-IL6-TNF+ cells. Lastly, the unstimulated B cell candidate biomarkers identified by the innate EN model included total IL10+ cells, GB-IL6-IL10+IL12-TNF- and GB-IL6+IL10-IL12-TNF+ cells. The values for recent and persistent QFT+ individuals for these variables were not significantly different, as indicated by Wilcoxon tests.

The final EN models built on the integrated variables had identical parameter values to the model for the adaptive data and consequently identified the same candidate biomarkers corresponding to an average AUROC value of 0.91.

To quantitatively evaluate whether a combination of adaptive and innate features improved the predictive performance of the model, a Likelihood Ratio Test (LRT) was used to compare the Logistic Regression (LR) model built to the 13 candidate biomarkers (three from the adaptive EN model and 10 from the innate model) to the LR model built to the three biomarkers from the adaptive EN only. The results indicated that a combination of the non-zero coefficients from both EN models did not significantly improve model fit (LRT *χ*^2^ = 6.09, p = 0.808). The LR model fitted to the adaptive biomarkers was therefore the preferred model.

Backwards variable selection on this model further identified *M*.*tb*-lysate-specific proportions of total Th1 cells expressing HLA-DR as a statistically redundant biomarker, since HLA-DR expression on either E6C10- or *M*.*tb*-lysate-specific T cells were highly correlated (Supplementary Figure 6).

The coefficients from the final LR model are shown in Table 1, model i. By exponentiating the coefficients, we can most easily interpret the coefficients in terms of the odds. Hence, holding all other variables fixed, for every one standardized unit increase in either HLA-DR or CD4+IL2+CD107-CD154-IFN-*γ*-TNF+ T cells in response to their specific stimuli, the odds of being a persistent QFT+ individual (the default class) decreases by 99% or 94% respectively. Accordingly, as the value of either one of these biomarkers increases, the odds that an individual was recently infected, i.e. QFT+, increases. The performance of this model was then assessed via an internal validation procedure and produced satisfactory results (average AUROC and Brier scores = 0.89 and 0.008 respectively).

**Table 1:**
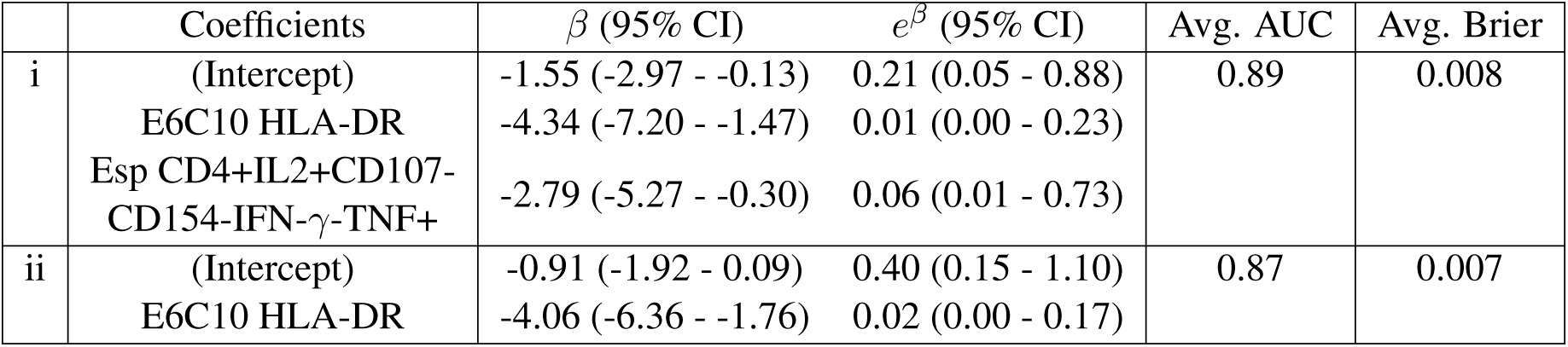
Model estimates and the average performance metrics after internal validation of the final LR model (i) and the LR model built to E6C10-specific total Th 1 cells expressing HLA-DR only (ii).

Candidate biomarkers from the innate dataset did not improve the adaptive model fit and these two variables from the adaptive dataset were sufficient to distinguish between the different stages of infection.

Since including the frequencies of Esp-specific IL2+CD107-CD154-IFN-*γ*-TNF+ CD4+ T cells as a predictor variable in the LR model statistically improved model fit (LRT *χ*^2^ = 12.76, p *<* 0.001) compared to E6C10-specific HLA-DR frequencies alone, we explored whether the successful discriminatory ability of this cell subset was dependent on the subset being negative for CD107, CD154 and IFN-*γ*. This was tested by comparing the predictive performance of an LR fitted to E6C10-specific HLA-DR and IL2+TNF+ CD4+ T cell when stimulated with Esp, regardless of CD107, CD154 and IFN-*γ* expression, to model i in Table 1. We found that the average AUROC value and hence predictive performance of this model was lower (average AU-ROC = 0.79) compared to model i. In addition, this model had a higher Akaike information criterion score [26] compared to model i (57 compared to 42), thus indicating a poorer fit to the data. The relative quality of the model was therefore dependent on the CD4+ T cell subset being negative for CD107, CD154 and IFN-*γ*.

The ability of E6C10-specific HLA-DR expression alone to distinguish between the different stages of *M*.*tb* infection was then assessed. The performance of this model and the model including Esp-specific IL2+CD107-CD154-IFN-*γ*-TNF+ CD4+ T cells were similar and equally high (average AUROC and Brier scores = 0.87 and 0.007 respectively) (Table 1, model ii).

### 2.4 Statistical validation

Using the same vast standardized and MFA-imputed dataset, a simple classification tree was built to all the 193 features in the dataset (Figure 3A). The tree identified two features from the set of all variables in the integrated dataset to best discriminate between recent and persistent QFT+ individuals. The best classifying feature in the dataset was the level of HLA-DR on total Th1 cells when stimulated with E6C10, followed by the frequency of Esp-specific IL2+CD107-CD154-IFN-*γ*-TNF+ CD4+ T cells. The split value for E6C10-specific HLA-DR was identified as −0.098 (Figure 3B). Seventeen observations had values greater than or equal to −0.098 for E6C10-specific HLA-DR expression levels and were assigned to leaf node 2, where all observations were correctly classified as recent QFT+. Otherwise, out of the seven observations in node 4, six were correctly classified as recent converters. Observations were assigned to this node if they had a value less than −0.098 for E6C10 HLA-DR but greater than −0.12 for the frequency of Esp-specific CD4+IL2+CD107-CD154-IFN-*γ*-TNF+ T cells (Figure 3C). Any observations that had values less than both these split values for each of the predictors were assigned to leaf node 5, which correctly classified 29 out of the total 30 persistent QFT+ individuals, but misclassified 6 recent QFT+ individuals. These decision rules identified by the tree resulted in 12% (7 out of 59) of the observations being misclassified.

The final RF model, after hyperparameter tuning, was then built such that 500 decision trees built to 500 random bootstrapped samples of the data made up the forest, a random subset of 25 out of the 176 features were considered at each split, and each tree built was allowed no more than 10 nodes from root to terminal node to avoid overfitting. The Gini Index was then used to measure variable importance. Among the 10 top variables with the largest mean decrease were nine variables from the adaptive dataset (Figure 3D). The E6C10-specific HLA-DR variable resulted in the largest mean decrease. One single variable from the innate dataset, total TNF production in unstimulated *γδ* T cells, was found to be the sixth most important.

After an internal model validation procedure, the average AUC for the final RF model was 0.84 and the average misclassification error was 0.12, precisely the misclassification error of the simple classification tree.

## 3 Discussion

This study applied regularized regression modelling approaches and machine learning algorithms to identify biomarkers that could distinguish individuals with remote or recent infection with *M*.*tb*, which is associated with higher risk of TB disease.

Data pre-processing steps were required to successfully integrate the datasets from the innate and adaptive immune responses. To overcome the high dimensionality of the integrated dataset, filtering methods were applied to each dataset separately. COMPASS [17] and MIMOSA [18] are very sensitive algorithms that have been developed to identify biologically meaningful combinations of cytokines produced by rare antigen-specific T cells, and significant responses over background, respectively. A combination of these methods was used to pre-filter the adaptive dataset. These methods assume background immune responses detected in unstimulated samples to be extremely low. This is not the case for most innate immune cells, which spontaneously produce variable levels of cytokines, even when cultured in absence of stimulation, that can be biologically meaningful. COMPASS and MIMOSA were thus not appropriate to pre-filter innate variables and we therefore developed our own filtering method to robustly identify biologically meaningful cell subsets from innate immune cells and DURT cells. The intrinsic biological variability between the two datasets was then accounted for by using vast scaling to standardize the raw values to a common scale, and missing values were successfully imputed using an MFA-based imputation method. We compared several imputation methods, to account for missing values, and the MFA-based method performed the best, in this dataset characterized by non-normally distributed data with missing rows. Because our focus in this aim was less on estimating model coefficients, but more on identifying predictive markers, instead of using Rubin’s rule [27] to take into account imputation variability, we rather repeated the MFA imputation for each CV run. Therefore, we were confident that the results found here were not a consequence of the imputation method used.

We first built an MTP-EN model, which applied differential penalties to the datasets in the integrated model. The results indicated that applying a greater penalty to the adaptive dataset (*κ <* 1) yielded comparatively worse AUROC values. However, when a smaller penalty was applied to this dataset (*κ >* 1), the predictive performances were identical to the model with no additional penalty (the standard EN model). In terms of computing power and predictive performance, there was insufficient evidence to justify building the MTP-EN model over the standard EN model for the integrated dataset. However, because a comparatively worse average AUROC value was yielded when a larger penalty was applied to the adaptive dataset, the MTP-EN results did demonstrate the importance of the features of the adaptive immune response in stratifying the two stages of *M*.*tb* infection.

The EN model built to the integrated dataset identified only non-zero coefficients from the adaptive dataset as important features. The candidate biomarkers identified from these models were the proportions of E6C10-specific Th1 cells expressing HLA-DR and the frequencies of Esp-specific IL2+CD107-CD154-IFN-*γ*-TNF+ CD4+ T cells. TNF and IL2 produced by CD4+ T cells are early response cytokines that both play an important role in the context of TB [28]. HLA-DR on the other hand is a cell surface receptor reflecting T cell activation. HLA-DR expression on *M*.*tb*-specific T cells is an excellent biomarker to distinguish individuals with (remote) *M*.*tb* infection from those with active TB disease, and to monitor antibiotic treatment response [29][30][31][32]. The robustness of HLA-DR expression as a biomarker to also distinguish recent from remote asymptomatic *M*.*tb* infection was confirmed in response to either E6C10 or *M*.*tb* lysate. We propose E6C10 to be a more appropriate stimulation for use in diagnostic tools, since it only includes *M*.*tb*-specific antigens (the same as in interferon gamma release assays), whereas *M*.*tb*-lysate contains a mix of different antigens that do cross-react with other mycobacteria and is therefore less specific.

The true effect of these two identified biomarkers on the probability of an individual being remotely *M*.*tb* infected (persistent QFT+ individuals) was estimated through a LR model. Higher standardized frequencies of both these biomarkers were associated with a larger probability, or odds, of an individual being recently infected (recent QFT+). This reflects the relationship that was seen in the raw data plots, where individuals recently infected with *M*.*tb* had significantly higher values of these features compared to remotely infected individuals. The performance of the LR model built to these two biomarkers was assessed via an internal validation procedure and, given the small sample size, was considered sufficiently high to justify further evaluation of these biomarkers.

A diagnostic test made up of a single biomarker would be simpler and more cost effective. Therefore, the ability of *M*.*tb*-specific T cell activation (HLA-DR expression) to successfully distinguish between the two stages of *M*.*tb* infection as a biomarker on its own was tested. Moving forward with HLA-DR as a single diagnostic measure was justified by substantial literature showing excellent performance of this biomarker to distinguish different stages of the TB spectrum [30][31][32][33][14], and the small number of markers necessary to measure this biomarker (as few as four [29]). Further, frequencies of the Esp-specific IL2+CD107-CD154-IFN-*γ*-TNF+ CD4+ T cells were extremely low (values range between 0 and 0.006), which is challenging to measure in a robust and reproducible way. Lastly, because the discriminatory ability of Esp-specific IL2+CD107-CD154-IFN-*γ*-TNF+ CD4+ T cells was in fact dependent the subset being negative for CD107, CD154 and IFN-*γ*, the flow cytometry antibody panel for a diagnostic test including all these markers would be complex. Although the final LR model included frequencies of Esp-specific IL2+CD107-CD154-IFN-*γ*-TNF+ CD4+ T cells as an additional biomarker of recent infection, further analyses showed that E6C10-specific HLA-DR expression alone is an equally strong single biomarker to distinguish recent from remote *M*.*tb* infection.

The top performance of *M*.*tb*-specific T cell activation over all other immune features as a biomarker of recent infection was further confirmed in both the simple decision tree and random forest model.

Lastly, in contrast to our hypothesis, variables from the innate dataset did not improve model fit and were unable to outperform the strongest candidate biomarkers from the adaptive dataset.

To our knowledge, this study includes the most comprehensive integrated evaluation of adaptive and innate immune responses induced by recent *M*.*tb* infection in humans published to date. Our results show that the innate immune responses were poor predictors of recent *M*.*tb* infection, and did not improve the performance of the integrated model. Based on the results reported here, the expression of HLA-DR on E6C10-specific T cells was the strongest candidate biomarker for recent *M*.*tb* infection, and its performance has now been validated in a separate test cohort [14]. This biomarker holds the potential to identify individuals at high risk of TB progression, who would benefit from preventive TB treatment. However, due to the small sample size in this study, further validation in a large independent cohort is required.

## 4 Materials and methods

### 4.1 Study design and participants

An epidemiological study was carried out from July 2005 through February 2009, in which healthy, 12 to 18-year-old adolescents were recruited from local high schools in the Worcester area, Western Cape, South Africa [11][12]. The study was approved by the University of Cape Town Human Research Ethics Committee (protocol references: 045/2005). Written assent from participating adolescents and written consent from their parents or legal guardians was obtained prior to the study start. Participants who tested human immunodeficiency virus (HIV) positive, were diagnosed with TB, or had any other acute or chronic medical diseases that resulted in hospitalization during the study period, were excluded from the study. Pregnant or lactating females were also excluded. Peripheral blood mononuclear cells (PBMCs) were collected at enrolment and at 6-monthly intervals during the 2-years of follow-up (termed months 0, 6, 12 and 18) when the QFT tests were performed to determine *M*.*tb* infection. The QFT tests were performed and interpreted according to the manufacturer’s instructions. Two cohorts of participants were defined based on their longitudinal QFT results and taking into account our proposed uncertainty zone to interpret quantitative values [13]: recent QFT converters (two consecutive QFT negative results, of which at least one is *<* 0.2 IU/mL, followed by consecutive two QFT positive results, of which at least one is *>* 0.7 IU/mL) and persistent QFT positives (QFT positive results *≥* 0.35 IU/mL at four consecutive visits) (Supplementary Figure 7). Raw QFT results and participant demographics have been described in detail elsewhere (training cohort in [14]).

### 4.2 Definition of recent and remote *M*.*tb* infection

Infection with *M*.*tb* likely occurred between the second and third sampling occasions in the recent QFT+ individuals, indicated by a QFT test conversion from negative to positive. After testing that there was no difference over time using Wilcoxon’s signed rank test, we calculated the median value of the two QFT positive time points for each variable in recent QFT+ individuals (n = 29 for the adaptive dataset and n = 16 for the innate dataset).

Time of *M*.*tb* infection was unknown in persistent QFT+ individuals (n = 30 for the adaptive dataset and n = 17 for the innate dataset). Since the Friedman’s test [15] did not reveal any significant changes over time, we included median values of each variable measured at all four QFT+ time points available as representative of remote *M*.*tb* infection (Supplementary Figure 7).

### 4.3 Immune measurements

Innate and adaptive effector responses were measured in stimulated PBMCs using flow cytometry (Supplementary Methods S1 and [16]). Five stimulations were used to induce *M*.*tb* and non-specific T cell responses in the adaptive dataset: peptide pools spanning E6C10; peptide pools spanning EspC, EspF and Rv2348c (collectively termed Esp), which are both are specific for *M*.*tb*; *M*.*tb*-lysate, which is a mixture of *M*.*tb*-specific antigens, some of which are cross-reactive with other mycobacteria; Staphylococcus Enterotoxin B (SEB), the positive control; or the cells were left unstimulated as negative control. This dataset consisted of 259 variables including a combination of 5 effector functions, namely interleukin-2 (IL2), CD107, CD154, IFN-*γ* and tumor necrosis factor (TNF) produced by CD4+ and CD8+ T cells upon stimulation. Combinations of the phenotypic markers CD45RA, CCR7, CD27, KLRG1, HLA-DR and CXCR3 were further measured on IFN-*γ*, IL2 or TNF producing T cells (total Th1). Effector responses were background subtracted (subtracting the frequencies detected in corresponding unstimulated samples from frequencies in stimulated samples), while the phenotypic markers were expressed as proportions of Th1 cells. Further, phenotypes were only measured in “responding” samples (Supplementary Method S3.2).

In the innate dataset, effector responses were measured in unstimulated PBMC or after stimulation with *M*.*tb*-lysate or *Escherichia coli* (*E. coli*), which served as the positive control. The innate dataset consisted of 304 variables, including a combination of 6 functions, Granzyme B (GB), IL6, IL10, IL12, IFN-*γ* and TNF produced by NK cells, B cells, monocytes, and DURT cells: mucosal associated invariant T (MAIT) cells, *γδ* T cells and NKT cells.

### 4.4 Data integration

Integration was performed by aligning each dataset according to participant ID, QFT status (positive or negative) and month of sample collection (months 0, 6, 12, and 18).

### 4.5 Data pre-filtering

Due to the high dimensionality of the dataset post-integration, we opted to pre-filter the dataset to identify and exclude biologically irrelevant cell subsets. For the adaptive dataset, we employed COMPASS (Combinatorial Polyfunctionality analysis of Antigen-Specific T cell Subsets) to filter the effector functions ([17]; Supplementary Method S2.1), while the phenotypic markers expressed on T cells were only measured in stimulated samples from responding individuals identified by MIMOSA (Mixture Models for Single Cell Assays [18]; Supplementary Method S2.2). Since COMPASS could not be used to filter the innate immune cells and DURT cells, which have high background (unstimulated) values, we designed a novel filtering method to identify biologically meaningful cell subsets from this dataset (Supplementary Method S2.3).

All analyses were performed on the pre-filtered dataset. An outline of the data pre-processing steps is provided in Figure 1A.

### 4.6 Data standardization

We employed variance stabilizing (vast) scaling to standardize the datasets. Vast scaling is achieved by multiplying the *Z*-score by a coefficient of variation (cv) as a scaling factor. Multiplying by the cv, which is the sample mean divided by the sample standard deviation of each variable, gives higher importance to those variables with small relative standard deviations. Vast scaling method aims to be robust and is typically used on variables that show small fluctuations [19].

### 4.7 Missing value imputation

Several imputation methods (Supplementary Table 2) were considered to meaningfully replace the missing values in the filtered and vast scaled dataset. The performance of each method was evaluated based on how well the imputed data could replicate the density of the vast scaled, incomplete data.

### 4.8 Standard elastic net model and elastic net model with multiple tuning parameters

A standard EN penalty is given by 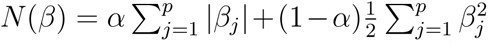, where 0 *≤ α ≤* 1 is;the weight given to the *L*_1_ penalty (the LASSO model) and 1 *− α* the weight to *L*_2_ penalty (ridge regression). The EN logistic regression model then aims to solve min_*β∈*ℝ_^*p*^ − *l*(*y, X*; *β*_0_, *β*) + *λN* (*β*) where *l*(*y, X*; *β*_0_, *β*) is the log-likelihood for a logistic regression model, *λ ≥* 0 is the shrinkage parameter, and the parameters *α* and *λ* are found via cross validation (CV). As the value of *λ* increases, the more coefficients of features are shrunk to zero, and their effect in the model is negligible.

The MTP-EN, proposed by Liu and colleagues in 2018 [7] for improving the predictive performance of a model fitted to an integrated dataset, extends the standard EN model by imposing separate penalties to the coefficients from different data types. The integrated dataset in this study was comprised of two different data types and the tuning parameters *λ*_1_ and *λ*_2_ were applied the coefficients from the adaptive and innate datasets separately. The MTP-EN model aims to solve the penalized regression problem given by

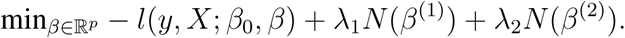

The model was built to the variables in the integrated dataset using the glmnet R package [20] via the “penalty.factor” argument. This allows a weighted EN penalty of the form

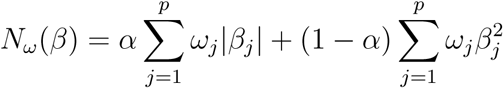

where *ω* is p-dimensional weight vector with 1 in the first *p*_1_=132 entries corresponding to the variables in the first dataset, and 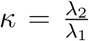 for the *p*_2_= 61 entries in the second dataset. Consequently, *λ* = *λ*_1_ controls the overall degree of shrinkage for both data types and *κ* controls the shrinkage of one data type relative to the other.

An outline of the workflow for the modeling portion of this study is summarized in Figure 1B. For each candidate weight parameter *κ ∈* [0.2, 1.8], 10-fold CV was used to tune optimal values for *λ* and *α* for a specific value of *κ*. The CV procedure was repeated 500 times for stable estimates and the area under the receiver operating curve (AUROC) was used as a measure of performance. The highest AUROC values after 10-fold CV were stored for each of the 500 repeats, and the performance of the MTP-EN model for each value of *κ* was reported as an average of the 500 AUROC values. A parameter value of *κ* = 1 is equivalent to a standard EN model, and so we could directly compare the performance of the standard EN model to MTP-EN models with varying penalties applied to each dataset. The result of this experiment was used to determine whether an MTP-EN or standard EN model would be the most suitable for the integrated dataset.

We further built two EN models using the glmnet package to the individual datasets separately. Parameters were tuned using the same CV protocol as the MTP-EN, and the average of the selected parameters across the 500 searches were thereafter defined as the “optimal” parameter values. Relevant candidate biomarkers (CB) for classifying *M*.*tb* infection were identified as features with non-zero coefficients in the final model, and predictive performances in terms of AUROC values of the models were then compared.

### 4.9 Logistic regression

The CBs identified from the innate and adaptive EN models were used to build LR models. One LR model was built using the biomarkers identified in the adaptive model, and another using a combination of the biomarkers identified from both the adaptive and innate data EN models. A LRT was used to assess whether adding the innate biomarkers to the LR model resulted in a statistically significant improvement in the fit of the model. Backwards variable selection was performed on the preferred LR model, as established by the LRT in the previous step, to identify the best subset of predictors and build the final LR model.

### 4.10 Tree-based machine learning algorithms

#### 4.10.1 Decision trees

A simple classification was built to all of the observations in the integrated dataset using the R package rpart [21]. The decision tree was used to visualize the relationship between the variables in the integrated dataset and assess feature importance in stratifying the recent from persistent QFT+ individuals.

#### 4.10.2 Random forest models

We built the RF model to our data using the randomForest R library [22], and tuned the model using 500×10-fold CV. Similar to the EN models, the “optimal” hyperparameters were taken as the average across the 500 repeats and used to build the final RF model. We then used the final RF model to identify the 10 most important features corresponding to the largest mean decrease in the Gini Index.

### 4.11 Internal validation

The predictive performance of the final LR model, and subsequently the set of biomarkers, as well as the final RF models was assessed via an internal validation procedure. We employed 10-fold CV repeated 500 times and used the AUROC and the Brier score [23] as performance metrics. Results reported are an average of the performance metrics across the 500 CV repeats.

For all instances in this study when CV was performed, the missing values in the dataset was imputed separately for the training and testing sets using MFA imputation. Therefore, the dataset was imputed several times to ensure that any results found were not just a consequence of the imputation method.

## Data Availability

Data will be available once the pre-print is published in a peer-reviewed journal.

## Author Summary

EN, MH, TJS raised funding and designed the study. CAMM, PS, CS, BM, TDR generated the data. TL, CAMM, VR, PS analyzed the data. TL, PS, EN and FL interpreted the results. TL, PS, EN and FL wrote the manuscript. All authors revised and approved the manuscript.

## Acknowledgments

We are thankful to the ACS study participants and their families; and the SATVI clinical and laboratory teams.

