## Supplementary Material for "Multidimensional analysis of immune response identified biomarkers of recent *Mycobacterium tuberculosis* infection"

ACS Study Team: Hassan Mahomed, Willem A. Hanekom, Fazlin Kafaar, Leslie Workman, Humphrey Mulenga, Ashley Veldsman, Rodney Ehrlich, Mzwandile Erasmus, Deborah Abrahams, Anthony Hawkridge, E. Jane Hughes, Sizulu Moyo, Sebastian Gelderbloem, Michele Tameris, Hennie Geldenhuys, Gregory Hussey.

#### 1 Supplementary Methods

##### 1.1 PBMC stimulation and staining protocol

To determine cytokine responses of innate and DURT cells (innate panel), cryopreserved PBMC were thawed, washed and rested for 2 hours in R10 media [RPMI 1640 (Gibco), 10% Fetal Bovine Serum (Gibco), 1% L-glutamine (Gibco) and 1% penicillin-streptomycin (Gibco)] prior to stimulation. Cells were then stimulated in R10 containing *M.tb*-lysate (H37Rv, 10 $\mu$g/mL, BEI Resources) to determine responses against mycobacterial antigens, heat killed Escherichia coli (E. coli, 107 bacilli per 1x106 cells, in house production) or left unstimulated (negative control). Cells were stimulated for a total for 6 hours at 37°C with 5% CO2. Brefeldin A (5$\mu$g/mL, Sigma Aldrich) and Monensin (2.5$\mu$g/mL, Sigma Aldrich) were added after the first 2 hours of stimulation and incubated for another 4 hours until harvest. After incubation, cells were detached from tubes using 2mM EDTA (Sigma Aldrich) in PBS (Lonza). Staining (Supplementary Table 1) for viability and surface markers was performed for 30 minutes at room temperature. Following surface staining, cells were washed, permeabilized and fixed (CytoFix/CytoPerm, BD Biosciences) for intra-cellular staining (ICS) of cytokines. ICS was performed for 30 minutes at room temperature. Prior to acquisition on a LSRII flow cytometer (BD Biosciences), cells were washed and fixed [1% paraformaldehyde (Kimix) PBS].

For the adaptive panel, PBMCs were processed and stained as described in [1].

#### 1.2 Data pre-filtering

##### 1.2.1 COMPASS

COMPASS [2] was used to pre-filter the adaptive dataset by removing binary cytokine combinations that were not biologically meaningful. The filtering protocol was applied to CD4+ and CD8+ T cells for each antigen specificity separately. A subset was classified as biologically meaningful if the number of observations with posterior probability values greater than 0.1, at any one of either month 0, 6, 12 or 18, was greater than 10 (one third of the number of participants in one cohort). The posterior probabilities are used to quantify the likelihood of detection of Ag-specific responses. For example, E6C10-specific CD4+ T cells with a joint expression of all five cytokines would be omitted from the final dataset because the number of observations with posterior probabilities greater than 0.1, at all sampling occasions, is less than 10 (Supplementary Figure 1).

##### 1.2.2 MIMOSA

We employed MIMOSA [3] to identify which individuals had a significant antigen-specific T cell response over background (unstimulated condition). Responding subjects are identified by testing whether the proportion of cytokine-producing cells in stimulated and unstimulated samples are different from each other. We defined as responders those individuals with a Th1 response in the stimulated samples that had a 3-fold change over unstimulated samples and a MIMOSA false discovery rate p-value less than or equal to 0.01. Since background expression cannot be subtracted for phenotypic markers, we measured these markers only in stimulated samples from responder individuals.

##### 1.2.3 Innate pre-filtering

COMPASS and MIMOSA have been designed to analyze antigen-specific T cell responses, with the assumption that background cytokine expression in unstimulated samples is generally very low. This is not the case for innate responses, where spontaneous cytokine production can occur even in absence of stimulation and could be biologically meaningful. To our knowledge, no similar computational tools exist that could handle variable and sometimes high background responses. In addition, we measured a variety of functional markers that could be expressed by multiple cell types, but we did not expect that all cell types would express all functional markers included in the panel (i.e. some measurable combinations are not biologically meaningful). Therefore, we established our own pre-filtering protocol for the innate dataset, summarized in Supplementary Figure 2.

We defined a threshold value that would identify whether a cell subset expressing different functional markers was detectable or not (i.e. biologically meaningful). Responses were considered as detectable if:

- the upper bound of the 95% confidence interval around the median, which was calculated across all samples, identified by bootstrapped methods, was non-zero, and

- one third of all samples have values greater than zero.

As a first step, this detection criterion was applied to the total cytokine variables. The goal was to identify which cytokines each cell type was able to produce in response to either *M.tb*-lysate or *E.coli* (the positive control for this dataset) stimulations. If the total cytokine variable was detectable in response to either *M.tb*-lysate or *E.coli*, then the variable would be retained. Otherwise, if the total cytokine variable was considered undetectable for both stimulations, then the variable was removed from the analysis of that cell type, including the binary functional combinations.

The second step focused on filtering the binary combinations of the different cytokines for each cell type. We first tested whether the binary subset was detectable when it was left unstimulated. If it was detectable, we further tested whether the *M.tb*-lysate stimulated version of this subset was significantly higher than when left unstimulated. If the lower bound of the 95% CI around the median for the *M.tb*-lysate sample was greater than the upper 95% CI of the median for the unstimulated version, the *M.tb*-lysate stimulated sample was kept in addition to the unstimulated sample. Otherwise, the *M.tb*-lysate stimulated sample was discharged and only values of the unstimulated version were kept. The rationale was that if the *M.tb*-lysate and unstimulated values were the same, then the biological responses were not different and hence it was unnecessary to keep both versions. If the unstimulated version of a cell subset was found to be undetectable, we tested whether the *M.tb*-lysate stimulated version was considered detectable. If it was, then the *M.tb*-lysate version was kept, otherwise we discarded both versions.

Lastly, for *M.tb* lysate-specific subsets that were preserved post-filtering, we performed background subtraction by subtracting the values measured in the unstimulated sample from the same set (i.e. same individual and time point). The exception for this was when the cell subset was Granzyme B positive. As Granzyme B is a cytotoxic molecule that is constantly present in cells and not only expressed after stimulation, it is not meaningful to perform background subtraction.

### 2 Supplementary Figures

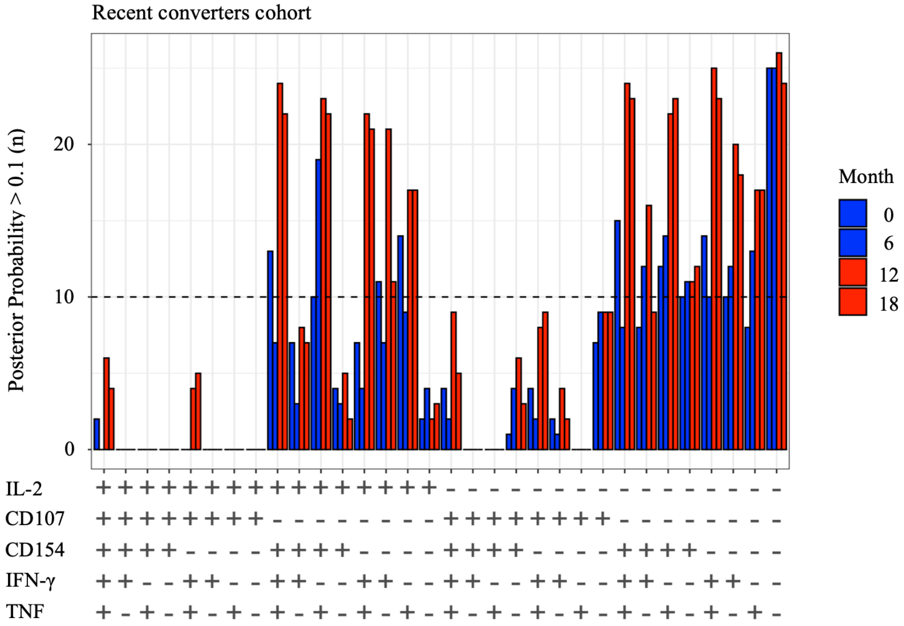


**Supplementary Figure 1: Filtering of the adaptive dataset.** Number of observations for CD4+ T cell counts stimulated with E6C10 in the recent QFT+ individuals that had posterior probabilities (calculated by COMPASS) greater than 0.1 for each binary combination, stratified according to month. A subset was classified as biologically meaningful if the number of observations with posterior probability values greater than 0.1, at one of either month 0, 6, 12 or 18, was greater than 10 (one third of the number of participants in one cohort).

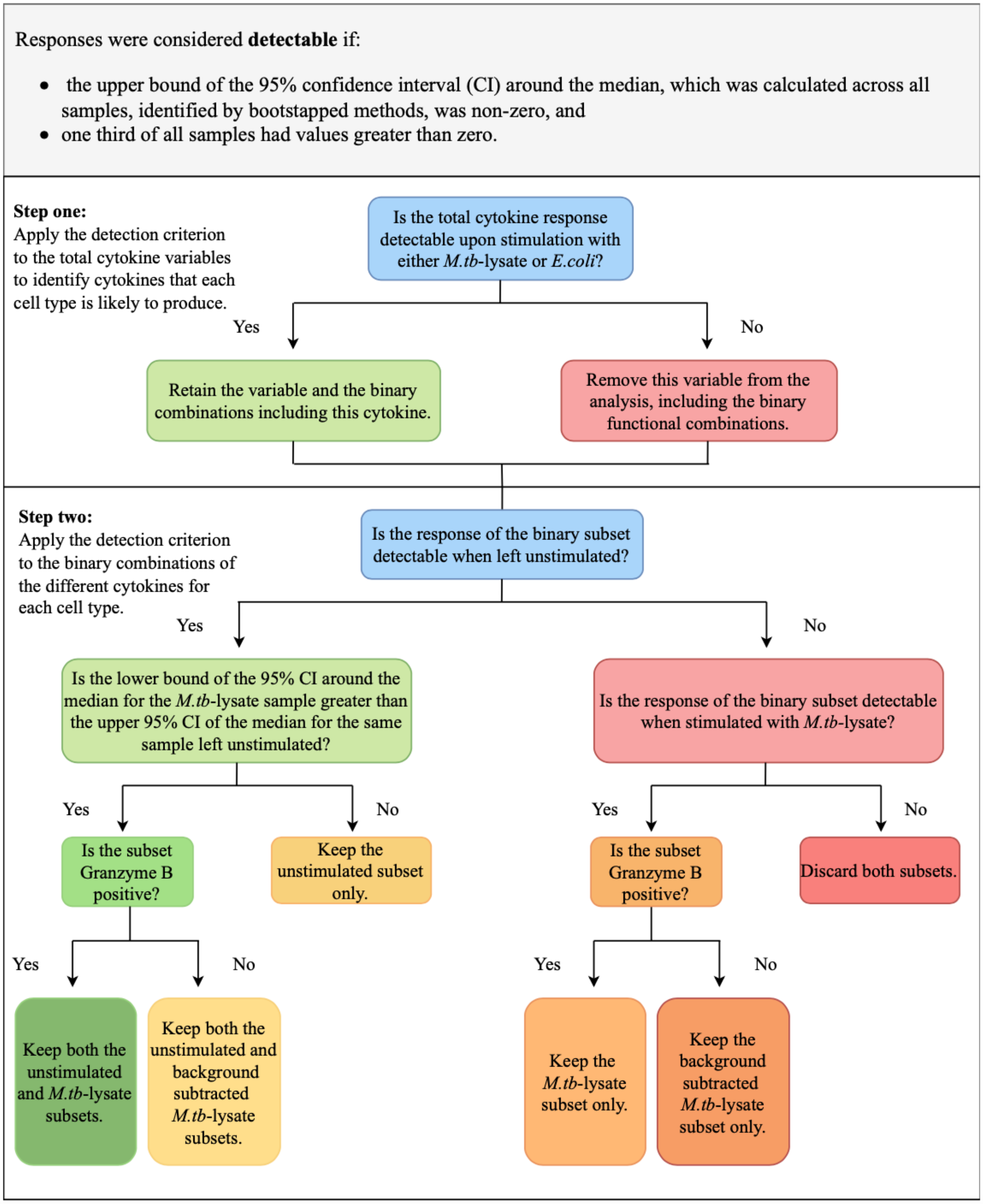


**Supplementary Figure 2: Flow chart for the innate data filtering.**

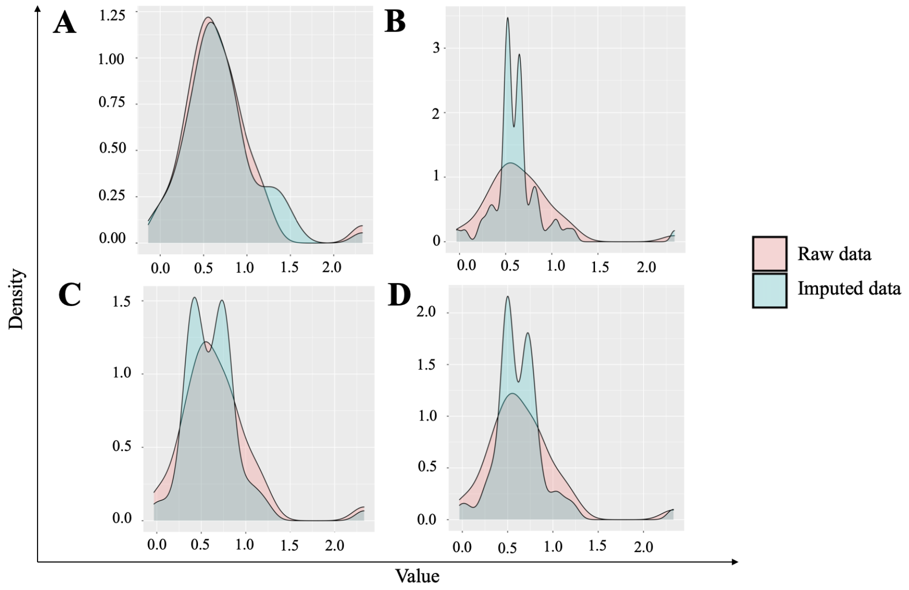


**Supplementary Figure 3: Data imputation methods.** The efficacy of each imputation method to capture the distribution of the raw frequencies of total IFN-$\gamma$ production in NKT cells stimulated with M.tb-lysate is shown as an example. The red lines are the raw data in each plot and the blue lines are (A) MFA, (B) column median, (C) $k$-nearest neighbours and (D) missForest imputed values.

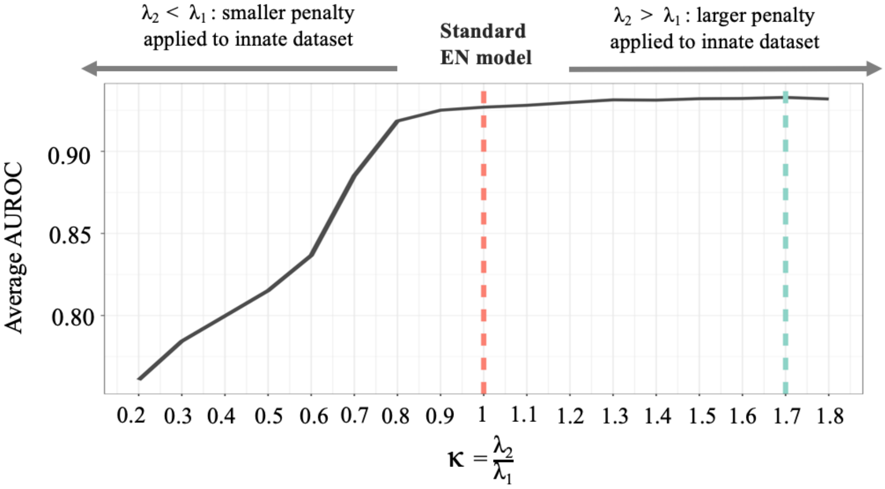


**Supplementary Figure 4: Performance of the MTP-EN model.** Average of 500 AUROC values is plotted as a function of $\kappa$, the ratio of the penalty parameter for the innate dataset relative to that for the adaptive dataset. When $\kappa < 1$ ($\lambda_2 < \lambda_1$) a smaller penalty is applied to the innate dataset, and when $\kappa > 1$ ($\lambda_2 > \lambda_1$) a larger penalty is applied to the innate dataset. A red dashed line is plotted at $\kappa$ = 1 ($\lambda_2 = \lambda_1$), which is equivalent to a standard EN model, and a blue line at the "optimal" $\kappa$ = 1.7, corresponding to the highest mean AUROC.

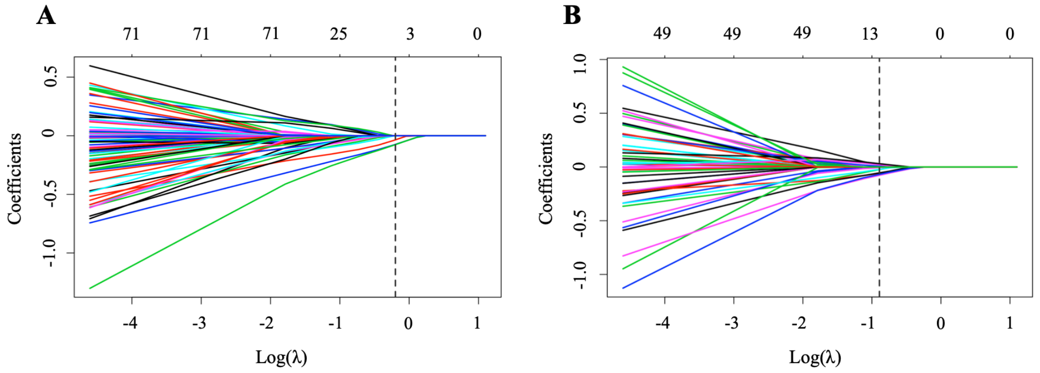


**Supplementary Figure 5: Coefficient paths for the final adaptive and innate EN model as a function of log($\lambda$).** Each line in the plots represents the coefficients of one variable for different values of $\lambda$, the overall shrinkage parameter in the EN model, from the respective datasets. An increasing value of $\lambda$ leads to the shrinkage of more regression coefficients and the number of non-zero coefficients for each $\lambda$ value are shown at the top of the figure. In the adaptive EN model (A) $\alpha$ was set to 0.21, where $\alpha \leq 1$ is the weight given to the L1 penalty and (1- $\alpha$) the weight to the L2 penalty. A dotted line is plotted at log($\lambda$) = -0.2 ($\lambda$ = 0.82), the optimal parameter values from the final adaptive EN model. At this point the number of non-zero coefficients are three and correspond to E6C10-specific or *M.tb*-lysate-specific HLA-DR expression on total Th1 cells and Esp-specific CD4+IL2+CD107-CD154-IFN-$\gamma$-TNF+ T cells. For the final innate EN (B) $\alpha$ was set to 0.21 and a dotted line is plotted at log($\lambda$) = -0.89 ($\lambda$ = 0.41) corresponding to 11 non-zero coefficients.

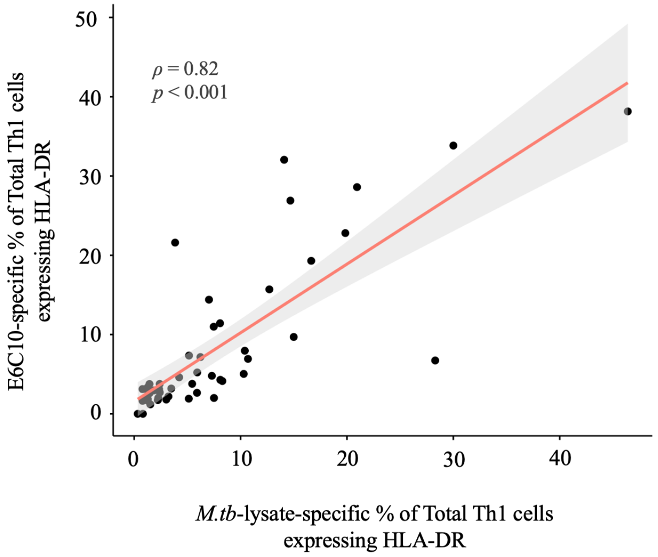


**Supplementary Figure 6: Correlation between M.tb-lysate (x-axis) and E6C10 (y-axis) stimulation on total Th1 cells expressing HLA-DR.** Spearman's non-parametric correlation coefficient and its associated p-value are superimposed onto the plot.

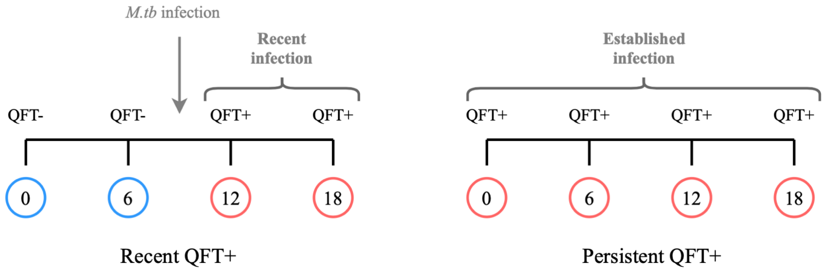


**Supplementary Figure 7: Cohort definition.**

### 3 Supplementary Tables

**Supplementary Table 1:** Antibodies for innate panel

| Marker | Fluorochrome | clone | Company |
|---|---|---|---|
| CD3 | BV786 | UCHT1 | BD |
| CD14 | PerCpeF710 | 61D3 | eBioscience |
| CD16 | AF488 | 3G8 | Biolegend |
| CD19 | BV711 | SJ25C1 | BD |
| CD26 | BV605 | M-A261 | BD |
| CD56 | BV50 | HCD56 | Biolegend |
| GB | BV510 | GB11 | BD |
| CD161 | PECy5 | DX12 | BD |
| gd TCR | BV421 | 11f2 | BD |
| IL-10 | PE-CF594 | JES3-19F1 | BD |
| IL-6 | PE | MQ2-13A5 | BD |
| IL-12 | APC | C11.5 | BD |
| IFN-$\gamma$ | AF700 | B27 | BD |
| TNF | PECy7 | MAb11 | BD |
| Live Dead | Near-IR | - | LifeTechnologies |

**Supplementary Table 2:** The various imputation methods that were tested.

| Method | Explanation | Author(s) | R package |
|---|---|---|---|
| missForest | A non-parametric imputation method that uses Random Forest models to predict the missing values and can handle mixed data | Stekhoven et al. (2012) [4] | missForest [5] |
| $k$ nearest neighbours (KNN) | An imputation method that employs the KNN algorithm to group variables with similar profiles. A weighted average of the $K$ nearest complete variables is taken and used to impute the missing values in the incomplete variables | Troyanskaya et al. (2001) [6] | DMwR [7] |
| Column median | Missing values were simply imputed with the column median of the incomplete variable | | |
| Multiple Factor Analysis (MFA) | Builds an MFA model on the incomplete dataset and uses the model to predict the missing values. | Husson et al. (2013) [8] | missMDA [9] |
